# Biological markers of brain network connectivity and pain sensitivity distinguish low coping from high coping Veterans with persistent post-traumatic headache

**DOI:** 10.1101/2024.09.16.24313761

**Authors:** Katrina S. Monroe, Dawn M. Schiehser, Aaron W. Parr, Alan N. Simmons, Chelsea C. Hays Weeks, Barbara A. Bailey, Bahar Shahidi

## Abstract

Headache is the most common type of pain following mild traumatic brain injury. Roughly half of those with persistent post-traumatic headache (PPTH) also report neck pain which is associated with greater severity and functional impact of headache. This observational cohort study aimed to identify biological phenotypes to help inform mechanism-based approaches in the management of PPTH with and without concomitant neck pain. Thirty-three military Veterans (mean (SD) = 37±16 years, 29 males) with PPTH completed a clinical assessment, quantitative sensory testing, and magnetic resonance imaging of the brain and cervical spine. Multidimensional phenotyping was performed using a Random Forest analysis and Partitioning Around Medoids (PAM) clustering of input features from three biologic domains: 1) resting state functional connectivity (rsFC) of the periaqueductal gray (PAG), 2) quality and size of cervical muscles, and 3) mechanical pain sensitivity and central modulation of pain. Two subgroups were distinguished by biological features that included forehead pressure pain threshold and rsFC between the PAG and selected nodes within the default mode, salience, and sensorimotor networks. Compared to the High Pain Coping group, the Low Pain Coping group exhibited higher pain-related anxiety (p=0.009), higher pain catastrophizing (p=0.004), lower pain self-efficacy (p=0.010), and greater headache-related disability (p=0.012). Findings suggest that greater functional connectivity of pain modulation networks involving the PAG combined with impairments in craniofacial pain sensitivity, but not cervical muscle health, distinguish a clinically important subgroup of individuals with PPTH who are less able to cope with pain and more severely impacted by headache.

## Introduction

Approximately 1.7 million individuals sustain a traumatic brain injury (TBI) requiring medical care each year in the United States, with nearly half of those hospitalized reporting residual disability one year later^1,2^. The majority of TBIs are classified as mild in severity and are characterized by post-concussive symptoms that can include headaches, musculoskeletal pain, dizziness, fatigue, sleep disturbances, and cognitive-emotional impairments^1-3^. Post-traumatic headache (PTH) is the most common type of pain reported among civilian^4^ and veteran^5^ populations with mild TBI, and nearly half of those who experience PTH report headache symptoms that persist 3 or more months after initial onset (i.e., persistent post-traumatic headache (PPTH))^6^. Approximately half of individuals with PPTH also report co-occurring neck pain^4,5,7^ which has been associated with greater severity and functional impact of headache^8^. Despite the high prevalence of PPTH, clinical management varies widely^9^ and treatment outcomes are often poor^10^. For example, individuals with PPTH frequently report that pharmacological interventions lack efficacy, and 87% of those receiving treatment in headache clinics or rehabilitation centers express dissatisfaction with their treatment status^7^.

Current treatment recommendations for PTH rely on guidelines for primary headache disorders due to a lack of evidence-based protocols for the management of secondary headaches following TBI^11,12^. This approach is likely insufficient to address the complex etiology of PTH, which can include neurologic, mechanical, metabolic, and/or psycho-emotional effects of head trauma^13-15^. PTH has a heterogeneous clinical presentation in which symptoms often resemble those of migraine but can also present as tension-type headache (TTH) or a mix of primary headache types^7,16,17^. Migraine and TTH are attributed to different pathophysiologic processes thought to involve sensitization of the trigeminovascular system^18^ or myofascial tissues in the craniocervical region^19^, respectively. In both types of primary headache, enhanced noxious signaling from sensitized peripheral tissues is thought to contribute to adaptations within the central nervous system that perpetuate spreading and chronification of pain^18,19^. Some have proposed that PPTH may also result from a neuropathic pain state involving peripheral sensitization of both trigeminal and cervical afferents coupled with nociplastic adaptions within the central nervous system^13,20^.

A growing body of evidence supports a role for nociplastic adaptations in pain sensitivity and functional connectivity of brain networks in the development and persistence of PTH. Several studies have used quantitative sensory testing to document localized mechanical hyperalgesia of craniocervical tissues in individuals with PPTH compared to headache-free controls with and without mild TBI, suggesting that PPTH involves regional sensitization of head and neck tissues^21-24^. Other studies have documented differences in connectivity within and between functional brain networks known to be involved in pain processing that distinguish PTH from other types of headache and pain-free controls^25,26^. Functional connectivity measures the strength of correlation between low frequency fluctuations in the blood oxygenation level-dependent (BOLD) functional magnetic resonance imaging (fMRI) signal that result from coordinated interactions between the spontaneous neural activity of anatomically distinct brain regions^27^. Longitudinal data have demonstrated that alterations in functional connectivity of the periaqueductal gray (PAG), a region within the brainstem that plays a critical role in the descending pain modulation network^28,29^, predicts the development of PPTH following TBI^30,31^. Structural evidence further suggests that reduced axonal connectivity of the PAG in individuals with mild TBI is associated with the clinical severity of post-traumatic pain^32^. Although existing studies have observed a wide variety of functional connectivity changes in PPTH^25,33^, some evidence supports attenuated connectivity of the PAG with both the default mode network (DMN)^30^ which regulates attention toward introspective thoughts and self-referential processing and the sensorimotor network (SMN)^31^ which discriminates and responds to sensory stimuli. The salience network (SN) is another network commonly studied in chronic pain disorders due to its role in coordinating responses to behaviorally relevant stimuli such as pain^29^. Alterations in functional connectivity of the SN have been associated with changes in emotional regulation following TBI^34^, however, the role of this network and its interaction with the PAG has not been studied in PPTH more specifically.

Brainstem regions including the PAG modulate activity of the trigeminal nucleus where there is convergence of afferent input from cerebrovascular, meningeal, and upper cervical structures^35,36^. It has been suggested that noxious afferent input resulting from direct mechanical injury to cervical tissues during head trauma may contribute to PTH^37-41^. This is supported by evidence that traumatic neck injuries such as whiplash associated disorder (WAD) share many of the distinguishing clinical features of post-concussive syndrome in patients with mild TBI, including headaches with concurrent neck pain that can persist for years after a traumatic injury^42,43^. Patients with WAD show consistent evidence of reduced size and quality of cervical muscles that are not present in the acute phase of a traumatic neck injury and are not observed in individuals who recover from WAD or in those with chronic idiopathic (i.e., non-traumatic) neck pain^44,45^. Prevalent among patients with chronic WAD, muscle atrophy and fatty infiltrates are hallmarks of impaired muscle health caused by chronic disuse, peripheral denervation, and/or degeneration following traumatic injury^45,46^. Although not directly investigated in the context of PPTH, these findings indicate that traumatized neck muscles exhibit maladaptive changes over time that may contribute to symptoms of persistent headache with concurrent neck pain.

Heterogeneity in the clinical presentation of PPTH and its pathophysiology is a major barrier for effective management of this disabling condition^47,48^. Identification of biologically based phenotypes is a critical step in the development of interventions that better target the mechanisms responsible for PPTH, particularly among those who experience high impact headaches with co-occurring neck pain. This study aimed to identify subgroups of individuals with PPTH who share common underlying impairments in functional connectivity of the PAG, cervical muscle health, pain sensitivity, and dynamic pain modulation. We hypothesized that a subgroup of individuals with impaired cervical muscle health and craniocervical hyperalgesia (peripheral impairments) combined with attenuated functional connectivity of the PAG and impaired pain modulation (central impairments) would report greater clinical severity of PPTH compared to individuals with impaired pain modulation in the absence of cervical muscle pathology.

## Materials and Methods

### Participants

A feasibility sample of 53 military Veterans with PPTH enrolled in this observational cohort study. MRI scans were collected for a subset of 38 participants. Five of these participants were excluded due to poor signal quality or technical issues during data collection. The final sample of 33 participants was powered to detect between-group differences having an effect size of 1.03 or larger with α=0.05 and power =0.80 (G*Power 3.1.9.7). Veterans with PPTH were recruited by convenience through a Veteran TBI Registry and clinic flyers and mailed advertisements at the Veterans Administration San Diego Healthcare System (VASDHS). Prospective participants were screened by telephone to determine eligibility. Criteria from the 2009 VA/Department of Defense (DOD) Clinical Practice Guidelines verified the occurrence and severity of TBI from electronic medical records^49^. We defined PPTH using diagnostic criteria from the International Classification of Headache Disorders (ICHD-III)^50^, including both acute- (within 7 days) and delayed-onset (up to 12 months) of headaches which continued to persist for 3 or more months after a traumatic head injury and were still present at the time of enrollment. To ensure that the study sample reflected the natural variation in co-occurring neck symptoms seen in the broader PPTH population, participants 18 to 60 years of age with a primary complaint of PPTH were enrolled regardless of the presence or severity of neck pain. Individuals with a history of severe TBI or persistent head or neck pain prior to the first-identified TBI were excluded. Additionally, we excluded those with widespread pain in multiple body regions other than the neck and head, and those reporting a lifetime history of chronic pain due to injuries other than TBI. Individuals with systemic (e.g., diabetes, lupus) or neurological (e.g., fibromyalgia, stroke) conditions potentially affecting sensation and those with major psychological conditions (e.g., bipolar disorder, psychosis) or with a current substance use disorder were also excluded. Due to well-documented effects of chronic opioid therapy on central pain processing^51^, those using opioids or other narcotic analgesics were excluded from participation and all other medications were recorded for analysis. Finally, pregnant females and individuals with standard contraindications for MRI were excluded for safety. These exclusion criteria were selected to minimize potential confounders while maintaining a valid representation of clinical characteristics that are prevalent among those with PPTH and may be important for phenotyping.

Eligible participants were scheduled for a single experimental session which included a semi-structured clinical interview followed by completion of demographic and clinical surveys, MRI scanning, and quantitative sensory testing by trained research personnel. Participants were asked to refrain from using alcohol or analgesic medications 24-hours prior to testing except when medically contraindicated. Written informed consent was obtained from all participants prior to study enrollment. The study was conducted in accordance with procedures approved by the VASDH) Institutional Review Board and Research and Development Committee.

### Clinical Interview and Survey Assessments

Participants completed a clinical interview with an examiner trained to administer the Boston Assessment of TBI – Lifetime (BAT-L)^52,53^ and a semi-structured headache history for TBI adapted from prior literature^16^. Following the clinical interview, participants completed sociodemographic and clinical surveys which were scored in the VA Research Electronic Data Capture (REDCap) platform. Self-reported demographics included age, height, weight, sex, race/ethnicity, education, and branch of military service.

The primary patient-reported outcome was headache-related disability measured by the Headache Impact Test (HIT-6). The HIT-6 has been shown to be valid and reliable in the assessment of primary headache disorders with a minimal clinically important change (MIC) of 5 points when anchored on either a 50% reduction in number of headache days or feeling much/very much improved following treatment^54,55^. The PROMIS Pain Intensity Scale^56^ (3a Adult – v2.0) was also used to assess pain intensity for headaches and neck pain separately. General psychological symptoms including depressed and anxious mood (PROMIS 8a Adult – v1.0 for Depression and Anxiety^57^, respectively), exposure to wartime stressors (Combat Exposure Scale^58^), and post-traumatic stress disorder (PTSD Checklist for DSM-5^59^; PCL-5) were assessed using validated measures. Raw PROMIS scores were converted to norm referenced T-scores, with values greater than 50 indicating more severe pain intensity relative to populations with at least minimal pain and more severe anxiety and depressive symptoms relative to the general adult population. Pain-related cognitive-emotional coping was evaluated using measures of anxiety and fear of pain (20-item Pain Anxiety Symptoms Scale^60,61^; PASS-20), pain catastrophizing (Pain Catastrophizing Scale^62^; PCS), and self-efficacy for managing pain (Pain Self Efficacy Questionnaire^63^; PSEQ).

### MRI Image Acquisition and Processing

Participants completed a 60-min MRI acquisition session to collect brain and cervical images. Scans were performed on a 3T GE Discovery MR750 (DV25) MRI System using a brain coil equipped with a cervical collar attachment (GE Signa Head Neck Spine Array (HNS) coil, SoundImaging Inc., no. 5341333). At the end of the data acquisition session, participants were asked to rate the severity of headache, neck pain, pain in other body regions, anxiousness, and sleepiness experienced while in the scanner using separate numeric rating scales.

### Brain Imaging

Participants were asked to rest quietly with eyes open during collection of anatomical and functional scans. Anatomical scans (MPRAGE) were collected with 208 sagittal slices (flip angle=8, TE= 2ms, TR=2500ms, TI=1060ms, acquisition matrix=320x320, reconstruction matrix=512x512, FOV=256mmx256mm, slice thickness=0.8mm, parallel imaging=2). The resting state acquisition (T2* EPI) was collected in three separate runs with 60 axial slices (flip angle=52, TE= 30ms, TR=800ms, acquisition matrix=104x104, FOV=208mmx208mm, slice thickness=2mm, multiband=6). The resting state seed-based analysis was performed using the standard afni resting state process pipeline through afni_proc.py (Appendix A). The primary region of interest (ROI) for the PAG was defined by a 3-mm radius sphere around MNI coordinates previously associated with altered PAG rsFC in primary migraine (2,-30,-6)^64^ and most closely aligned with coordinates identifying the dorsolateral PAG in prior studies^65^. Based on prior literature^66-68^, extraction ROIs were selected within the *default mode network* (DMN: medial prefrontal cortex (mPFC), posterior cingulate cortex/precuneus (PCC), subgenual anterior cingulate cortex (sgACC), lateral temporal cortex (LTC), inferior parietal cortex(IPC)), *salience network* (SN: temporoparietal junction (TPJ), anterior insula (aINS), dorsolateral prefrontal cortex (DLPFC), middle cingulate cortex (MCC)), and *sensorimotor network* (SMN: primary motor cortex (M1), primary sensory cortex (S1), supplementary motor area (SMA), thalamus, posterior insula (pINS), cerebellum).

### Cervical Imaging

Anatomical images of the cervical spine were obtained in supine using a high-resolution 3D iterative decomposition of water and fat with echo asymmetry and least squares estimation (IDEAL) sequence^69^ to quantify fat and lean muscle. Scans were acquired from the C1 to T1 vertebral levels according to the following parameters: TR=1974, TE=16, FoV=25.6, acquisition and reconstruction matrix 256x256, and 1mm^3^ voxel size. Bilateral cervical muscle (longus colli and capitus combined, cervical multifidus, obliquus capitus superior, and rectus capitus posterior major) volumes were extracted from 3D anatomical datasets using previously documented, semi-automated segmentation routines for spine muscles^70-72^. Whole muscle volumes were measured to account for postural and regional differences in muscle structure throughout the cervical spine across subjects. The fraction of fat within the muscle volume was calculated from 3D IDEAL images based on isolation of the independent contributions of water (S_w_) and fat (S_F_) to the total MR signal. Fat fraction (FF) was then quantified using the following equation^73^: 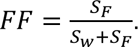 Whole muscle volume and FF values did not significantly differ across right and left sides of the body, therefore, individual values for each marker of cervical muscle health were averaged across sides for each muscle. Cervical MRI image processing was conducted by an assessor who was blind to the clinical characteristics and group status of study participants.

### Quantitative Sensory Testing (QST)

#### Pain Sensitivity

A trained examiner who was blind to imaging outcomes quantified pain sensitivity using a hand-held digital algometer (AlgoMed; Medoc Advanced Medical Systems, Israel) to assess pressure pain threshold (PPT) at four regional test sites on the head and neck (forehead, temporalis muscle, cervical spine, upper trapezius) and one remote test site on the volar forearm. Test sites were selected based on prior studies of PTH^22,74^. To minimize accessory muscle activity during testing, participants sat upright in a chair with the back and arms fully supported and the position of the head stabilized by a second examiner. Pressure stimuli were applied manually with a 1-cm^2^ rubber-tip algometer at a rate of 30 kPa/s^22,74^ using real-time force feedback visible only to the examiner. Participants were instructed to press a response button at the exact moment the pressure sensation from the algometer started to feel uncomfortable or slightly painful; the pressure at this moment was recorded and the mean of 2 trials at each test site (90-sec interstimulus interval (ISI)) was recorded as the PPT. Sites were tested in random order, and individual trials were repeated if pain intensity was rated greater than 2 on a numeric pain rating scale (NPRS)^75,76^ ranging from 0 (no pain) to 10 (worst pain imaginable). PPT values did not significantly differ across right and left sides of the body for muscles tested bilaterally, therefore, individual values were averaged across sides for each test site.

#### Dynamic Pain Modulation

*Temporal summation* (TS), quantified by the increase in pain intensity evoked by repeated application of a constant noxious stimulus, is considered a psychophysical correlate of centrally mediated wind-up in humans^77^. The present study adapted a protocol with established reliability for assessing mechanical TS in healthy adults^78^. Briefly, 10 consecutive pressure stimuli (application rate = 100kPa/sec, hold time = 1-sec hold, ISI = 1-sec) were applied to the right volar forearm using a handheld pressure algometer at a constant intensity equal to 1.5x the PPT assessed as described above for each participant. Two trains of 10 stimuli were separated by a 2-min rest break. Pain intensity was assessed during the first, fifth, and tenth trials of each train with a verbal numeric pain rating score (NPRS). Mechanical TS was quantified as the difference between the NPRS from the first trial and the highest score from subsequent trials within each train. TS values were averaged across both trains, with positive values indicating greater TS.

*Conditioned Pain Modulation* (CPM) refers to the change in pain intensity experienced at a primary test site when a second noxious stimulus is applied to a remote body region; in humans, this test is thought to reflect the efficiency of diffuse noxious inhibitory controls, a widespread reduction in afferent nociceptive signaling modulated by serotonergic spino-brainstem-spinal pathways^79^. We assessed CPM as the change in PPT (phasic test stimulus) for the right temporalis muscle immediately after submerging the left hand to wrist level in an 8°C circulating water bath (cold pressor test; tonic conditioning stimulus) compared to an identical control condition using tepid water to account for the non-thermal sensory effects of hand immersion. A sequential paradigm^80^ was used in which PPT was assessed twice (ISI = 15-sec) for the temporalis muscle^81^ immediately upon withdrawing the hand from the water bath in each temperature condition. The tepid condition was performed first to avoid carry over effects from the cold pressor response^82^. Normalized CPM was calculated as: CPM (%) = (mean PPT_TEPID_ – mean PPT_COLD)_ / mean PPT_TEPID_) * 100, such that negative values of CPM indicated more efficient endogenous pain inhibition^80^. Measurement reliability has been found to be excellent using the cold pressor test as a conditioning stimulus for CPM^83^.

### Statistical Analyses

#### Classification of Subgroups

Multidimensional phenotyping of the PTH cohort was performed with a Random Forest (RF) analysis and Partitioning Around Medoids (PAM) clustering of input features from 3 biologic domains: 1) brain network connectivity with the PAG, 2) cervical muscle health, and 3) quantitative sensory tests of pain sensitivity and modulation. RF is a machine learning approach that can be used to identify data features that are most important for classifying individuals with a heterogenous clinical condition into subgroups who share similar distinguishing characteristics. The RF is used as both a supervised and unsupervised learning method. This analysis has previously been used to identify subtypes of primary migraine^84^ and is preferred to other analytic approaches using multiple features because it is less affected by intercorrelated variables.^85^ The RF procedure generates an ensemble of loosely correlated decision trees using bootstrapped samples of the original data with approximately one-third of the data remaining out-of-sample for each tree used to calculate misclassification (out-of-bag error). The magnitude of the decrease in accuracy when a variable is excluded from the prediction of subgroup membership indicates the importance of that variable. This is determined by permuting each variable one at a time in the out-of-sample data.

An initial unsupervised RF analysis was conducted by generating a proximity matrix that included all variables across all three measurement domains (46 input features). The number of trees grown was 10000 and for every split in a tree, the RF selected a random subset of 7 predictor variables. Clustering with this proximity matrix and the PAM algorithm identified k=2 PTH subgroups having the largest average silhouette coefficient ^86^ plotted across a range of possible subgroup numbers. PAM is a clustering algorithm that is more robust to noise and outliers as compared to the more commonly used k-means algorithm. Subgroups were visualized in two dimensions using a multidimensional scaling (MDS) plot to display the proximity between data points in multiple dimensions reflecting non-linear combinations of variables included in the analysis.

To reduce the number of data features and prevent overfitting, a supervised RF analysis was then used to identify which predictor variables were statistically important (p<0.10) by ranking variables based on their importance for discriminating between the two subgroups. Variable importance was quantified by the mean decrease in accuracy, which reflects the extent to which model accuracy for identifying subgroups would decrease if the variable were removed from the analysis. To reduce error from extraneous features in the classification model, a final supervised RF analysis including only the statistically important variables was used to assign subgroup membership prior to assessing model accuracy. Error for identifying each subgroup and overall out-of-bag error for distinguishing subgroups with the final RF algorithm were calculated. Analyses to classify subgroups were performed with randomForest, rfPermute, and cluster packages in R version 4.2.0.^87^

#### Comparison of Subgroup Characteristics

Normality was assessed for all variables with the Shapiro-Wilks test and descriptive statistics were calculated as mean (SD) for parametric distributions and median (IQR) for nonparametric distributions. Differences in sociodemographic and clinical characteristics were compared between PPTH subgroups using independent sample t-tests for approximately normal data. Mann Whitney U was used as a nonparametric test and categorical variables were compared with Fisher Exact tests. Biologic features found to be important in differentiating PPTH subgroups were also compared using similar methods. Clinical correlates of biologic features were assessed by Pearson or Spearman Rank correlations using the HIT-6 as the primary clinical outcome for headache-related disability. Analyses used to compare subgroup characteristics and assess correlations with clinical outcomes were performed in SPSS version 28.0 using a significance criterion of α=0.05.

## Results

Sociodemographic and TBI characteristics for the full PPTH cohort (N=33) and subgroups identified by classification of biologic features are provided in **Table 1**. Consistent with other Veteran cohorts, participants were predominantly high school or college educated males ranging from 23 to 55 years of age. The sample was racially and ethnically diverse and represented all branches of military service, with the majority being Non-Hispanic Caucasian Navy Veterans. Eighty-eight percent of the sample experienced more than one TBI during their lifetime. The most significant TBI occurred between 1.5 to 32 years prior to enrollment, was most frequently attributed to a blunt head injury, and was classified as mild in severity for all participants. There was wide variation in the lifetime cumulative severity of TBI (BAT-L score range = 2 to 12 points). Sociodemographic and injury characteristics did not differ between subgroups (**Table 1**). There were also no significant differences in symptom severity on the day of testing for headache (median (IQR) NPRS points = 2(4) vs. 0(3), p=0.235), neck pain (2(3) vs. 2(4), p=0.928), other body pain (2(3) vs. 1(3), p=0.842), anxiousness (2(4) vs. 0(3), p=0.181), or sleepiness (7(7) vs. 7(5), p=0.986) for Low Pain Coping vs. High Pain Coping groups, respectively.

**Table 1.** Sociodemographic and Injury Characteristics for Persistent Post-Traumatic Headache Cohort.

|  | Full Cohort<br>(N=33) | Low Pain<br>Coping<br>(N=20) | High Pain<br>Coping<br>(N=13) | *p*-<br>value |
| --- | --- | --- | --- | --- |
| **Sociodemographic Characteristics** |  |  |  |  |
| Age (years)* | 37.0 (16.5) | 37.5 (16.3) | 37.0 (7.5) | 0.758 |
| Body Mass Index (kg/m<sup>2</sup>) | 29.9 (3.9) | 29.5 (4.2) | 30.4 (3.5) | 0.751 |
| Sex |  |  |  |  |
| Female | 4 (12) | 3 (15) | 1 (8) | 0.638 |
| Male | 29 (88) | 17 (85) | 12 (92) |  |
| Ethnicity |  |  |  |  |
| Not Hispanic | 23 (70) | 14 (70) | 9 (69) | 0.963 |
| Hispanic | 10 (30) | 6 (30) | 4 (31) |  |
| Race |  |  |  |  |
| White | 23 (70) | 13 (65) | 10 (77) | 0.914 |
| Asian/Pacific Islander | 5 (15) | 3 (15) | 2 (15) |  |
| Black | 4 (12) | 3 (15) | 1 (8) |  |
| Multiracial | 1 (3) | 1 (5) | 0 (0) |  |
| Education (years) | 14.7 (1.7) | 14.5 (1.6) | 15.1 (1.8) | 0.338 |
| Branch of Military Service |  |  |  |  |
| Navy | 17 (52) | 13 (65) | 4 (31) | 0.224 |
| Marine | 8 (24) | 4 (20) | 4 (31) |  |
| Army | 2 (6) | 1 (5) | 1 (8) |  |
| Air Force | 2 (6) | 0 (0) | 2 (15) |  |
| Coast Guard/National Guard | 2 (6) | 1 (5) | 1 (8) |  |
| Not reported | 2 (6) | 1 (5) | 1 (8) |  |
| **TBI Characteristics** |  |  |  |  |
| BAT-L score (0-60 points)* | 6.0 (5.0) | 6.5 (5.5) | 4.0 (3.0) | 0.235 |
| Number of lifetime TBIs | 5.7 (6.3) | 6.3 (6.9) | 4.9 (5.5) | 0.529 |
| Time since most significant TBI (months)* | 148.0 (111.5) | 154.0 (160.0) | 148.0 (63.0) | 0.703 |
| Grade of most significant TBI |  |  |  |  |
| Mild - Grade I | 6 (18) | 4 (20) | 2 (15) | 0.944 |
| Mild - Grade II | 22 (67) | 13 (65) | 9 (70) |  |
| Mild - Grade III | 5 (15) | 3 (15) | 2 (15) |  |
| Mechanism of most significant TBI |  |  |  |  |
| Blunt only | 27 (82) | 17 (85) | 10 (77) | 0.621 |
| Blast only | 5 (15) | 2 (10) | 3 (23) |  |
| Blast with secondary blunt | 1 (3) | 1 (5) | 0 (0) |  |
| Combat Related TBI |  |  |  |  |
| Yes | 14 (42) | 8 (40) | 6 (46) | 0.727 |
| No | 19 (58) | 12 (60) | 7 (54) |  |

Values are number (%) or mean (SD) unless otherwise noted as \*median (IQR)
TBI = Traumatic Brain Injury, BAT-L = Boston Assessment of Traumatic Brain Injury

### Subgroup Classifications

Two subgroups of individuals with PPTH were identified by the RF analysis and PAM clustering as shown by the MDS plot in **Figure 1**. These two groups were labeled ‘High Pain Coping’ and ‘Low Pain Coping’ based on the distinguishing clinical characteristics described below. The overall out-of-bag error for group classification was 6.1%, with less error in the classification of Low Pain Coping (0.0% classification error) compared to High Pain Coping (15.4% classification error) individuals. **Figure 2** shows the biologic features that best distinguished subgroups in the final supervised RF analysis. Features are shown in order of importance based on the mean decrease in accuracy (MDA) for each variable in the overall model, with MDA values plotted separately for each group. Important biologic features (p<0.10) included rsFC between the PAG and selected nodes within the default mode network (PCC/precuneus), salience network (MCC, TPJ, DLPFC), and sensorimotor network (SMA, M1, thalamus, S1, cerebellum). Only one quantitative sensory test (forehead PPT) was found to be important in distinguishing PPTH subgroups. Notably, no measures of cervical muscle health or dynamic pain modulation were important in the classification model.

**Figure 1.**
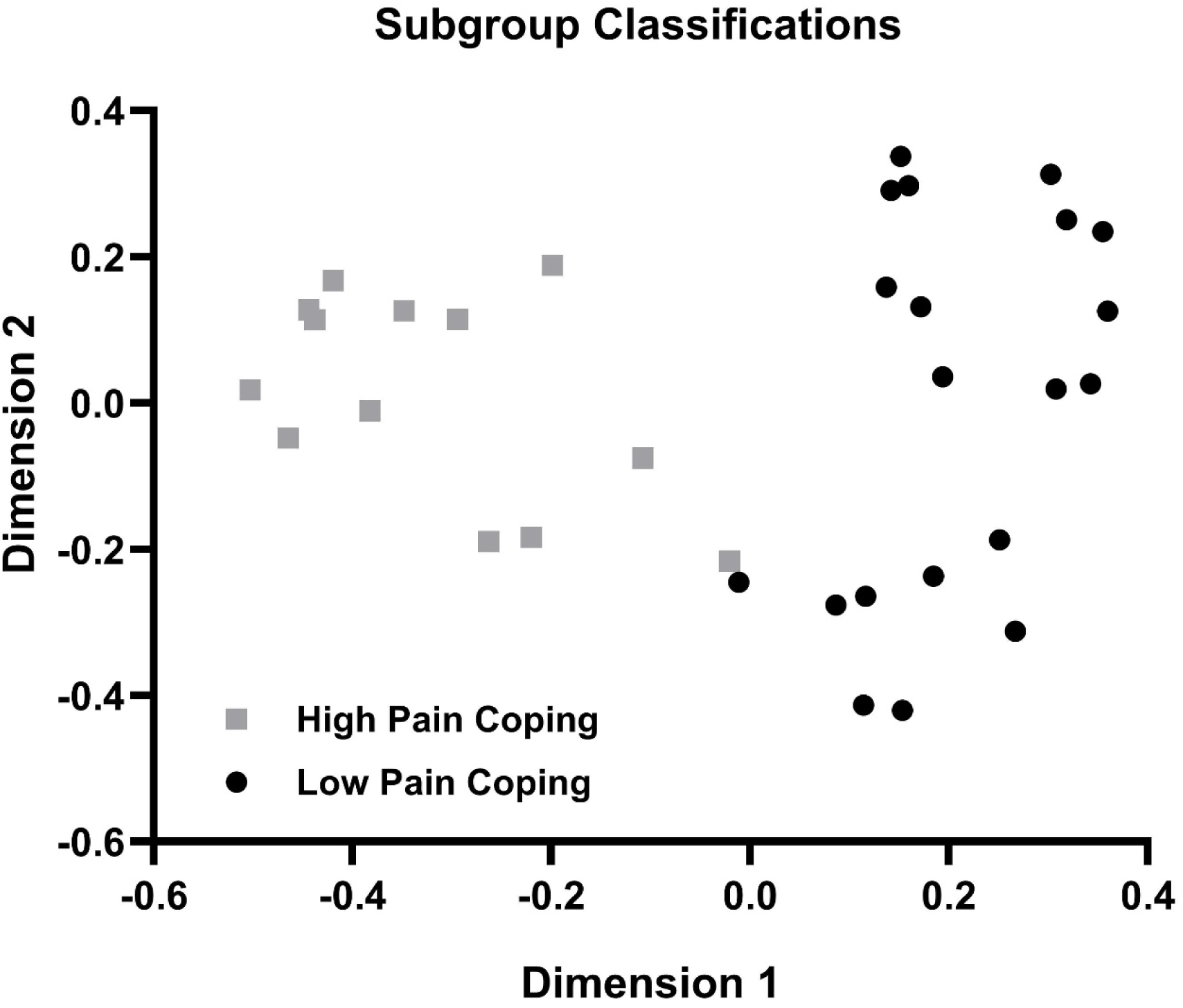
Multidimensional Scaling Plot of Subgroup Assignments for Persistent Post-Traumatic Headache Phenotypes. Two subgroups of individuals with persistent post-traumatic headache were identified using a Random Forest analysis and Partitioning Around the Medoids clustering of input features from 3 biologic domains: 1) resting state functional connectivity of the periaqueductal gray, 2) quality and size of cervical muscles, and 3) mechanical sensitivity and modulation of pain. Distinct clustering of participants assigned to High Pain Coping (gray squares, N=13) and Low Pain Coping (black circles, N=20) subgroups is visualized in two dimensions using multidimensional scaling to reflect non-linear combinations of features included in the analysis.

**Figure 2.**
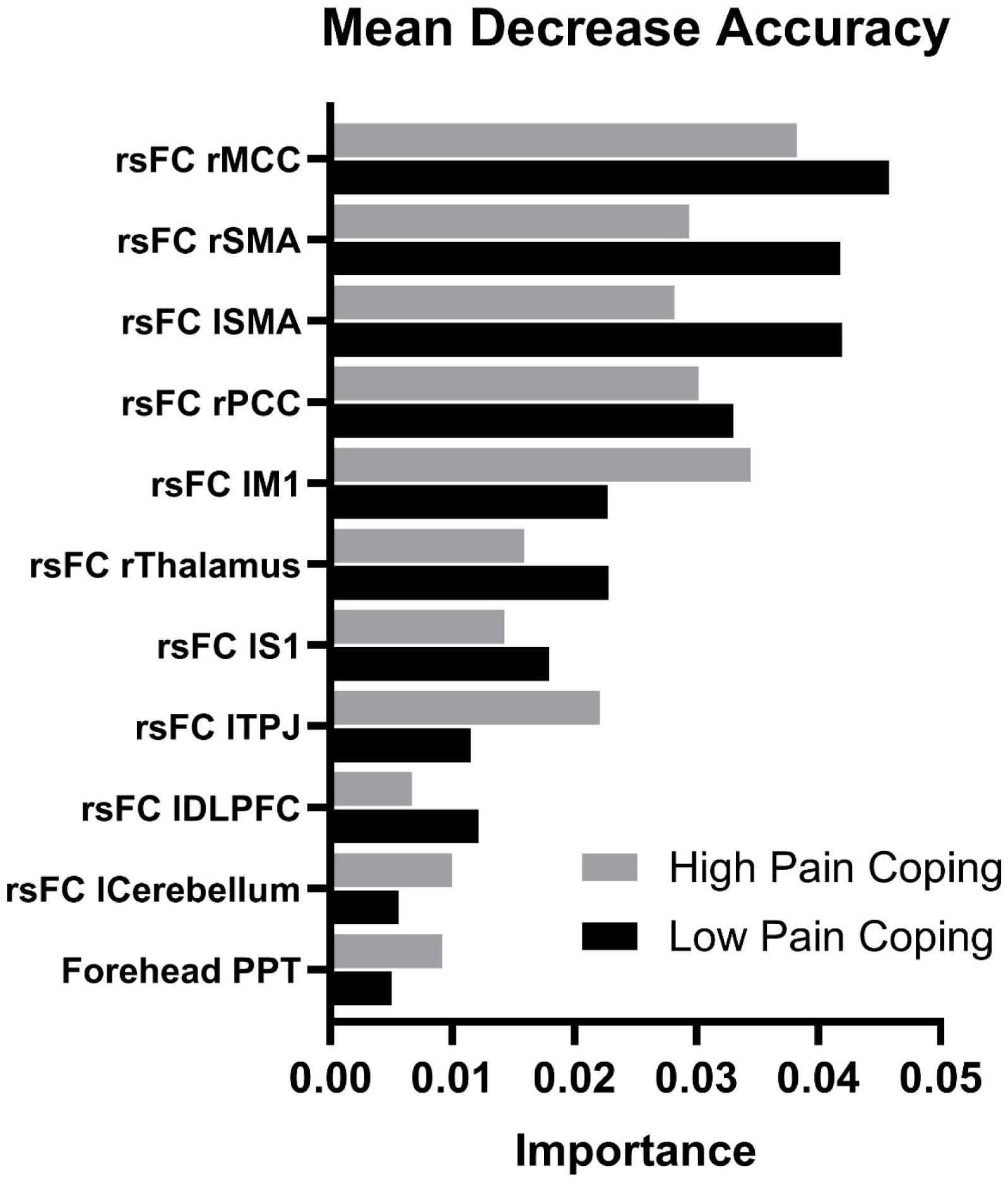
Mean Decrease in Accuracy (MDA) for Most Important Features Differentiating Persistent Post-Traumatic Headache Phenotypes. Significant features are shown in order of importance based on the MDA for each variable in the overall classification model. MDA values are plotted separately for High Pain Coping (gray bars, N=13) and Low Pain Coping (black bars, N=20) subgroups. Significant features included resting state functional connectivity of the periaqueductal gray with other brain regions and mechanical pain sensitivity at the forehead. Abbreviations: rsFC=resting state functional connectivity, r=right, l=left, MCC= middle cingulate cortex, SMA= supplementary motor area, PCC= posterior cingulate cortex/precuneus, M1= primary motor cortex, S1= primary sensory cortex, TPJ= temporoparietal junction, DLPFC= dorsolateral prefrontal cortex, PPT=pressure pain threshold.

### Subgroup Differences in Important Biologic Features

**Table 2** reports subgroup differences in mean (SD) Fisher z_r_ values reflecting the strength of PAG rsFC for all brain fMRI features found to be important in differentiating PPTH phenotypes. All features were significantly different between Low Pain Coping and High Pain Coping subgroups (p≤0.002). On average, PAG activity was positively correlated with the activity of all other network nodes of interest for the Low Pain Coping group whereas the High Pain Coping group showed anticorrelated activity.

**Table 2.**
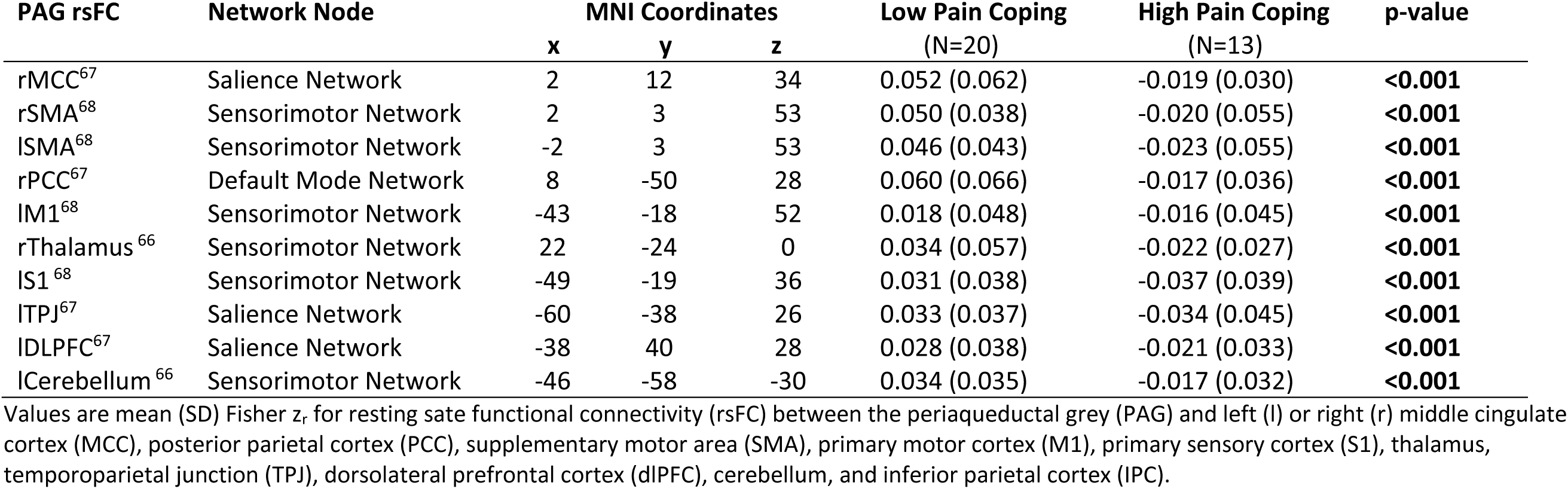
Subgroup Differences in Periaqueductal Gray Resting State Functional Connectivity for Most Important Features Differentiating Persistent Post-Traumatic Headache Phenotypes.

Subgroup differences in median (IQR) pain sensitivity (PPT) for regional and remote test sites are illustrated in **Figure 3**. The Low Pain Coping group had significantly greater regional pain sensitivity (lower PPT) at the forehead (p=0.048), consistent with the role of this feature as an important variable. PPT also tended to be lower for the Low Pain Coping group at other regional test sites, although these differences did not reach significance and were not considered important distinguishing features (temple PPT, p=0.080; neck PPT, p=0.080; shoulder PPT, p=0.281). There were no group differences in pain sensitivity assessed remotely at the forearm (p=0.501).

**Figure 3.**
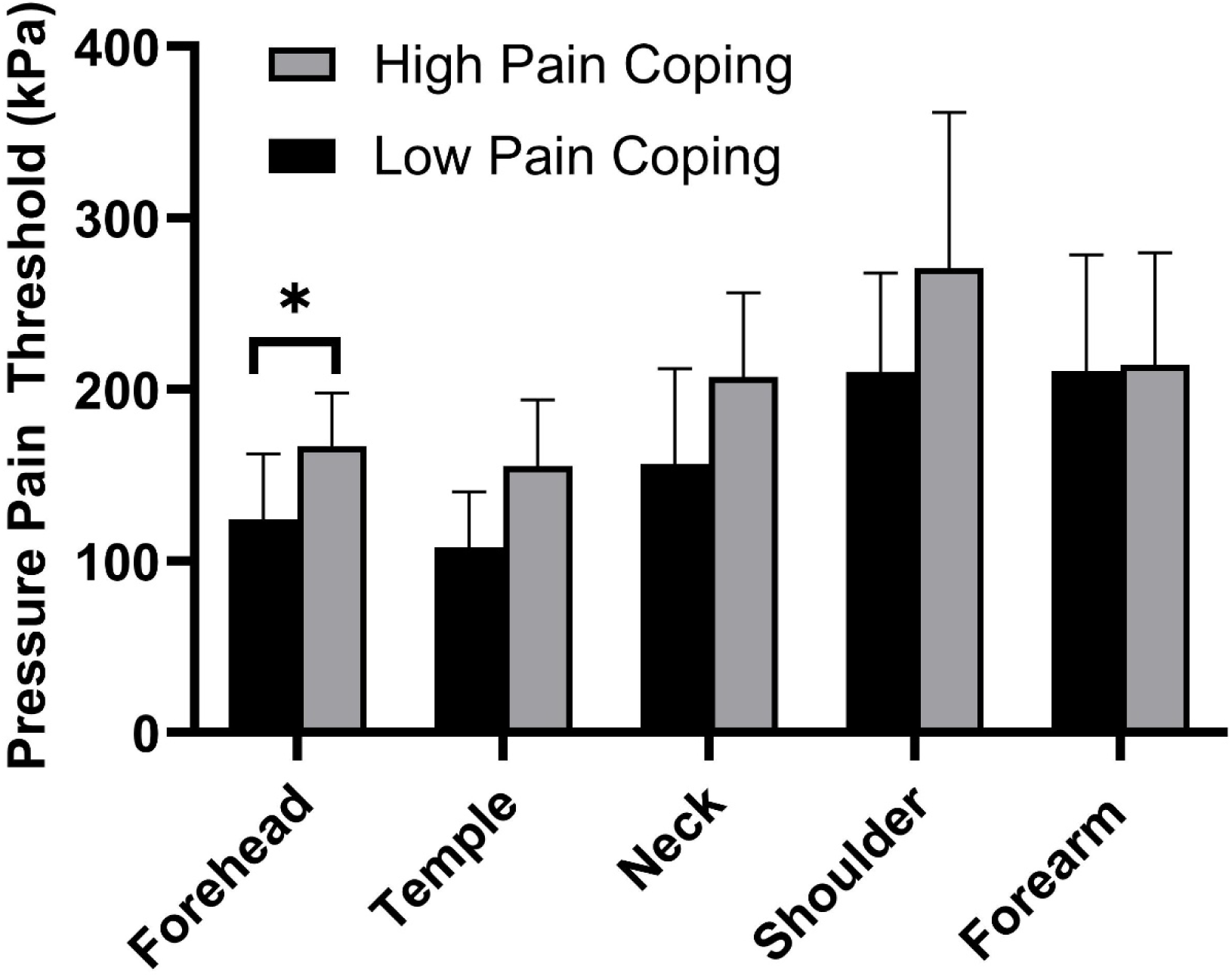
Comparison of Mechanical Pain Sensitivity for Persistent Post-Traumatic Headache Phenotypes. Bars show median (IQR) for pressure pain threshold (PPT) assessed at four local test sites in the craniocervical region and one remote test site at the forearm in High Pain Coping (gray bars, N=13) and Low Pain Coping (black bars, N=20) subgroups. There was a trend for lower PPT (i.e., higher pain sensitivity) in the group with low pain coping at all craniocervical test sites, with a significant difference between groups at the forehead (*p<0.05).

### Subgroup Differences in Clinical Characteristics

**Table 3** compares psychological and clinical characteristics of the two PPTH subgroups that were identified from biologic features. Clinically, these groups were distinguished by significantly greater pain-related anxiety (PASS, p=0.009), higher pain catastrophizing (PCS, p=0.004), lower pain self-efficacy (PSEQ, p=0.010), and more severe headache-related disability (HIT-6, p=0.012) for the Low Pain Coping group compared to the High Pain Coping group. Notably, differences in pain-related cognitive-emotional coping and headache disability were observed in the absence of group differences in more general cognitive-emotional symptoms (anxiety, depression, combat stress, post-traumatic stress) or the type, intensity, and frequency of headache and neck pain. Compared to the High Pain Coping group, a greater proportion of individuals in the Low Pain Coping group reported taking one or more prescription medications (62% v. 100%, p=0.005). The proportion of individuals using anti-inflammatory medications was higher (70% v. 23%, p=0.013) whereas use of cardiovascular medications was lower (10% v. 46%, p=0.035) in the Low Pain Coping group. Low and High Pain Coping groups did not differ in the prevalence of psychoactive or analgesic medication use (45% v. 46%, p=0.948) or in the use of preventive/abortive medication for headache (25% v. 0%, p=0.131).

**Table 3.** Comparison of Psychological and Clinical Characteristics for Persistent Post-Traumatic Headache Subgroups Identified from Biologic Features.

|  | Low Coping<br>(N=20) | High Coping<br>(N=13) | P-Value |
| --- | --- | --- | --- |
| **Psychological Characteristics** |  |  |  |
| PROMIS Depression (T-score) | 56.9 (7.3) | 54.2 (9.1) | 0.364 |
| PROMIS Anxiety (T-score) | 61.3 (5.7) | 58.2 (5.4) | 0.125 |
| Combat Exposure Scale (0-41 points)* | 18.0 (21.8) | 12.0 (18.5) | 0.650 |
| PTSD Checklist for DSM-5 (0-80 points) | 39.6 (16.4) | 30.0 (10.7) | 0.073 |
| Pain Anxiety Symptoms Scale (0-100 points)* | 40.5 (32.3) | 14.0 (29.0) | **0.009** |
| Pain Catastrophizing Scale (0-52 points)* | 18.0 (12.0) | 8.0 (12.0) | **0.004** |
| Pain Self-Efficacy Questionnaire (0-60 points) | 32.2 (9.4) | 44.1 (15.6) | **0.010** |
| **Clinical Characteristics** |  |  |  |
| Headache Impact Test-6 (36-78 points) | 63.2 (4.4) | 56.1 (8.4) | **0.012** |
| Headache Type |  |  |  |
| Migraine-like | 4 (20) | 1 (7) | 0.327 |
| TTH-like | 0 (0) | 1 (7) |  |
| Mixed | 16 (80) | 11 (86) |  |
| PROMIS Pain Intensity – Headache (T-score) | 59.9 (6.8) | 56.6 (7.3) | 0.186 |
| Headache frequency |  |  |  |
| Once a month or less | 0 (0) | 2 (15) | 0.428 |
| Once a week | 6 (30) | 3 (23) |  |
| Several times a week | 11 (55) | 6 (47) |  |
| Daily or Constant | 3 (15) | 2 (15) |  |
| PROMIS Pain Intensity – Neck Pain (T-score) | 59.3 (10.3) | 58.6 (5.7) | 0.837 |
| Neck pain frequency |  |  |  |
| Once a month or less | 4 (20) | 1 (8) | 0.320 |
| Once a week | 5 (25) | 1 (8) |  |
| Several times a week | 2 (10) | 4 (31) |  |
| Daily or Constant | 9 (45) | 7 (53) |  |

Values are number (%) or mean (SD) unless otherwise noted as \*median (IQR)
PROMIS=Patient-Reported Outcome Measurement Information System; PTSD=Post-traumatic stress disorder; DSM = Diagnostic and Statistical Manual of Mental Disorders, TTH=Tension Type Headache

### Correlates of Headache-related Disability

Headache-related disability (HIT-6) was moderately correlated with rsFC between the PAG and the cerebellum (r=0.51, p=0.003), IPC (r=0.42, p=0.016) and TPJ (r=0.41, p=0.019), indicating that higher levels of disability were associated with greater connectivity between each of these brain regions and the PAG. HIT-6 scores were also inversely correlated with regional but not remote pain sensitivity (forehead PPT, ρ=-0.45; p=0.008; temple PPT, ρ=-0.44; p=0.011; neck PPT, ρ=-0.47; p=0.005; shoulder PPT, ρ=-0.38; p=0.032; forearm PPT, ρ=-0.30; p=0.094), indicating that higher levels of disability were associated with greater craniocervical pain sensitivity (i.e., lower PPT). Finally, there was a moderate inverse correlation between HIT-6 and longus colli muscle volume (r=-0.41, p=0.018), indicating that higher levels of disability were associated with less muscle volume. No other indices of dynamic pain modulation (CPM, TS), muscle size (volume), or muscle quality (FF) were significantly correlated with headache disability.

## Discussion

This is the first study to perform multidimensional phenotyping using biologic indicators of central and peripheral impairments in the same cohort of individuals with PPTH. Contrary to expectation, subgroups were predominantly differentiated by functional connectivity of the PAG with other brain networks known to be involved in pain processing and not by differences in the size, quality, or mechanical sensitivity of cervical muscles. Compared to high coping individuals with PPTH, low coping individuals had greater rsFC between the PAG and selected nodes within the DMN, SMN, and SN, along with heightened mechanical pain sensitivity in the forehead. Clinically, individuals with low pain coping reported greater pain-related anxiety, higher pain catastrophizing, and reduced self-efficacy for managing pain. These individuals also reported greater impacts of headache on daily functioning. Group differences in pain-related coping and disability were observed despite no differences in general mood or the type, frequency, and intensity of head and neck pain. Collectively, these findings suggest that a subgroup of individuals with high levels of disability and more maladaptive coping styles may tonically engage top-down pain modulation pathways while at rest, yet greater engagement of the PAG by higher brain centers appears insufficient to reduce heightened pain sensitivity of craniofacial tissues in these individuals.

### Functional Connectivity of the Periaqueductal Grey in Headache

The PAG is critical for homeostatic regulation of salient body functions. This includes modulation of pain through opioid- and non-opioid mediated pathways engaged by distinct anatomical subdivisions within the PAG.^88^ Prior research has demonstrated that the PAG is susceptible to axonal injury following TBI^32,33^ and exhibits altered functional connectivity with key regions of the DMN and SMN that correlate with acute symptom severity and predict the development of persistent post-traumatic head and neck pain^30,31^. Our results extend these findings by demonstrating that differences in rsFC between the PAG and both the DMN (PCC/precuneus) and SMN (S1, M1, SMA, thalamus, and cerebellum) are important for distinguishing subgroups with clinically meaningful differences in headache disability in the chronic stage of PTH. Interestingly, a recent tractography study using whole-brain graph analysis also identified the PCC/precuneus (implicated in emotional salience and multisensory integration) as a primary pathologic hub in episodic migraine^89^. These findings suggest that dysfunction of DMN connectivity may be a common mechanism underlying both PPTH and chronic migraine.

We also examined connectivity between the descending pain modulation network and the SN which helps detect and coordinate responses to salient stimuli by switching from activation of the DMN during self-referential mind wandering to activation of the executive control network during goal-oriented attention to potentially noxious stimuli^29^. For the first time, we show that rsFC between the PAG and SN (MCC, TPJ, and DLPFC) is important in distinguishing PPTH phenotypes, with PAG-MCC connectivity emerging as the most important feature in the classification model. The MCC is activated by nociceptive stimuli from the thalamus and has direct projections to the PAG^90^. This region is involved in the cognitive-emotional control of behavioral responses to pain, particularly fear-evoked nocifensive behaviors^91,92^. The TPJ and DLPFC both function to integrate top-down contextual knowledge with bottom-up sensory information when coordinating behavioral responses to pain^93^.

The prominent role of PAG connectivity with brain regions involved in cognitive-emotional processing of pain is well aligned with the clinical phenotypes of PPTH subgroups identified by these features. Specifically, PPTH subgroups differed in clinical measures of pain-related cognitive-emotional coping (i.e., pain-related anxiety, pain catastrophizing, and pain self-efficacy) with no appreciable differences in more general affect or the type, frequency, or intensity of headache and neck pain. Moreover, high coping individuals with less pain-related anxiety and catastrophizing and greater self-efficacy for managing pain reported less impact of headache on their daily lives with a clinically meaningful difference in the magnitude of HIT-6 scores^54^. Notably, PAG connectivity with key nodes within the DMN (IPC) and SN (TPJ) were correlated with headache disability, consistent with a prior study of DMN connectivity in acute PTH^30^. Together, these findings suggest that intrinsic connectivity of the PAG with brain networks underlying cognitive-emotional regulation of pain plays an important role in behavioral coping and disability resulting from PPTH. This is aligned with a primary role of the dorsolateral PAG in coping strategies that involve active defense behaviors in response to perceived threats^65,88^. Behavioral^94^ and neuromodulatory^95^ interventions shown to modulate these brain networks and improve symptoms in other chronic pain disorders may hold promise for enhancing clinical outcomes in individuals with PPTH and warrant further investigation.

Interestingly, the PAG showed uniformly anticorrelated activity with all brain networks in high coping individuals, whereas those with low pain coping showed positively correlated activity. This finding was unexpected given prior observations that attenuated functional connectivity between the PAG and DMN has been associated with worse clinical outcomes following acute PTH^30,31^. Previous reports in healthy adults have described both positive and anticorrelated connectivity of the PAG with multiple brain regions^65,96^, including the majority of those identified as important features of PPTH in the present study. While there is general consensus that positively correlated activity reflects functionally coordinated activity of anatomically remote brain regions, the interpretation of anticorrelated activity is less clear. Some evidence suggests that negative correlations may be an artifact of global signal removal^97^, whereas other studies indicate that anticorrelated activity is a neural phenomenon reflecting opposing functions of distinct networks that remain detectable with alternate methods of preprocessing^93,98,99^. From this perspective, it is possible that positively correlated network activity in individuals with low pain coping reflects greater intrinsic engagement of the descending pain network by the DMN and SMN at rest, as well as by the SN when transitioning attention toward salient stimuli such as ongoing or spontaneous pain. Conversely, anticorrelated activity in individuals with high pain coping could indicate disengagement of the PAG by these networks while at rest. While greater engagement of the PAG may be beneficial during recovery from acute PTH^30,31^, overreliance on pain modulation networks in the presence of poor cognitive-emotional functioning may be maladaptive in the chronic stage of headache. Given that the same preprocessing steps were used for all participants, it seems unlikely that differences in correlated and anticorrelated network activity were solely caused by processing artifacts. Additional studies are needed to clarify the interpretation of differences in the relative magnitude and directionality of correlated BOLD signals in the acute and chronic stages of PTH.

### Pain Sensitivity and Dynamic Pain Modulation in Headache

Systematic reviews of mechanical pain sensitivity in primary headache show consistent evidence of reduced craniocervical PPT in individuals with migraine and TTH compared to healthy controls^100,101^. A fewer number of investigations report similar findings for individuals with PTH^21-24,74^. The largest of these studies^21^ (N=200) found significantly reduced PPT values for individuals with PPTH compared to healthy controls for the temporalis (159 vs. 207 kPa) and upper trapezius (259 vs. 313 kPa) muscles, respectively. PPT measured at the same test sites in the present cohort were comparable to values reported by Ashina et al.^21^ for individuals with PPTH and 25-40% lower than pain-free controls. These observations are consistent with prior reports suggesting that heightened sensitivity of craniocervical tissues to noxious mechanical stimuli is a characteristic feature of headache disorders, including PPTH. In contrast to localized differences in forehead PPT, we found no difference in pain sensitivity at the forearm between low and high coping PPTH subgroups, suggesting that heightened sensitization in lower functioning individuals may be localized to the trigeminal cervical complex. This finding aligns with the notable absence of remote mechanical hyperalgesia in prior studies comparing PTH to headache-free controls with and without TBI^22-24,74^ and suggests a lack of widespread central sensitization extending beyond the region of localized injury.

Few studies have evaluated dynamic measures of central pain modulation in PTH. Using methods similar to the present study, Carey et al.^74^ found less efficient CPM but no difference in mechanical TS among individuals with acute TBI compared to healthy controls. Consistent with findings from our PPTH cohort, deficits in CPM were not associated with headache severity during the acute phase of TBI. In contrast, Defrin et al.^23^ found deficiencies in CPM relative to pain-free controls with and without TBI which were correlated with headache intensity in a PPTH population. Findings on CPM in primary headache are also mixed^102^, with some evidence suggesting that CPM declines more rapidly with repeated exposure to noxious stimuli in migraineurs despite being initially similar to healthy controls^103^. While speculative, these findings could indicate more rapid depletion of endogenous mechanisms for central inhibition of pain in chronic headache disorders characterized by frequent episodes of pain. Waning efficiency of CPM with repeated engagement of the descending pain modulation network might explain why resting connectivity of higher brain centers to the PAG was found to be important in differentiating PPTH phenotypes in the present study, whereas the response to a single CPM challenge was not. Additional studies of dynamic pain modulation are needed to clarify the role of central inhibition and facilitation of pain in both primary and secondary headache disorders.

### Cervical Tissue Health

Despite suggestions that noxious afferent input from injured or deconditioned cervical tissues might contribute to sensitization of the trigeminal cervical complex in headache disorders^37^, few imaging studies have examined cervical tissue health in relation to PPTH. One retrospective analysis of patients treated for head trauma found that reduced size of the rectus capitus posterior minor was associated with the severity of acute PTH and predicted recovery time for post-concussive symptoms^41^. Although our results cannot determine the extent to which cervical muscles may be impaired relative to headache-free controls, we found little evidence to support a major role for muscle size or quality on the severity of PPTH in our cohort. Reduced volume of the longus colli muscle was moderately associated with greater headache disability, but no such associations were observed for muscle volume or fat fraction for other cervical muscles examined in this study. Additionally, no indices of cervical muscle health were important in distinguishing PPTH phenotypes when considered alongside other indices of brain connectivity and pain sensitivity. These findings were unexpected, given strong evidence supporting a role for cervical muscle injury and deconditioning in chronic WAD^44,45,104^ which shares many clinical features of mild TBI including comorbid headache and neck pain^40^. Prospective longitudinal studies similar to those for WAD are needed to clarify the role of cervical tissue injury and subsequent changes in muscle health on recovery profiles for PTH and neck pain following TBI.

### Study Limitations and Future Directions

Several important limitations should be addressed in future investigations. First, we explored a large number of candidate biomarkers in a small exploratory cohort of Veterans with PPTH. To minimize overfitting, we used a multistep process of variable reduction with a machine learning approach that is robust to correlations within the feature set and has been shown to perform with high accuracy even with small sample sizes (N<50)^105^. However, our findings may not generalize to civilian or athlete populations, or to individuals with acute PTH. Well established biologic and clinical sex differences in headache^106^ could also affect the biologic features important for distinguishing subgroups of PPTH among women. Therefore, findings from our classification model require independent external validation in a larger and more heterogeneous sample. Input features were selected *a priori* based on prior literature and did not include many features that might also be important in differentiating biologic phenotypes of PPTH. For example, mechanical injury of non-muscular cervical structures (e.g., ligament, disc) have been suggested as peripheral sources of neck pain in both headache and WAD, albeit with limited evidence^107,108^. Our brain imaging analysis focused on functional connectivity of the PAG as a primary region of interest due to its role in central pain modulation and evidence of changes in axonal connectivity following TBI. Future studies could examine phenotypes using network connectivity or task-related activation of other brain regions previously associated with PPTH^26,109^.

## Conclusion

Findings suggest that greater functional connectivity of pain modulation networks involving the PAG combined with impairments in craniofacial pain sensitivity, but not cervical muscle health, distinguish a clinically important subgroup of individuals with PPTH who are less able to cope with pain and more severely impacted by headache. These individuals may tonically engage top-down pain modulation pathways while at rest, yet greater engagement of the PAG by higher brain centers appears insufficient to reduce heightened pain sensitivity of craniofacial tissues. Screening and targeted interventions to improve cognitive-emotional pain coping may help improve outcomes for this subgroup of individuals with PPTH.

## Data Availability

All data produced in the present study are available upon reasonable request to the corresponding author.

## CRediT Authorship

**Monroe KS**: Conceptualization, Methodology, Formal Analysis, Investigation, Data Curation, Writing – Original Draft, Visualization, Supervision, Project Administration, Funding Acquisition. **Schiehser DM**: Conceptualization, Methodology, Data Curation, Writing – Review and Editing, Supervision, Project Administration, Funding Acquisition. **Parr A**: Investigation, Data Curation, Writing – Review and Editing. **Simmons AN**: Conceptualization, Methodology, Formal Analysis, Review and Editing, Visualization. **Bailey B**: Formal Analysis, Review and Editing, Visualization**. Shahidi B:** Conceptualization, Methodology, Formal Analysis, Investigation, Data Curation, Writing – Review and Editing, Visualization, Supervision, Project Administration, Funding Acquisition.

## Author COI

The authors declare no conflicts of interest.

## Funding

This work was supported by the National Institute of Neurological Disorders and Stroke under NIH award R21NS109852 and the VA Medical Center of San Diego Center of Excellence for Stress and Mental Health (CESAMH). The content is solely the responsibility of the authors and does not necessarily represent the official views of the NIH or CESAMH. The funder played no role in the design, conduct, or reporting of this study.

## Notes

### Competing Interest Statement

The authors have declared no competing interest.

### Funding Statement

This study was funded by the National Institute of Neurological Disorders and Stroke [R21NS109852] and the VA Medical Center of San Diego Center of Excellence for Stress and Mental Health.

### Author Declarations

The ethics committee/IRB and Research and Development Committee of the Veteran's Administration of San Diego Health System gave ethical approval for this work.

## REFERENCES

1. Corrigan JD, Selassie AW, Orman JA. The epidemiology of traumatic brain injury. J Head Trauma Rehabil 2010;25(2):72–80, doi:10.1097/HTR.0b013e3181ccc8b4

2. Theeler B, Lucas S, Riechers RG, 2nd, et al. Post-traumatic headaches in civilians and military personnel: a comparative, clinical review. Headache 2013;53(6):881–900, doi:10.1111/head.12123

3. Schwab K, Terrio HP, Brenner LA, et al. Epidemiology and prognosis of mild traumatic brain injury in returning soldiers: A cohort study. Neurology 2017;88(16):1571–1579, doi:10.1212/WNL.0000000000003839

4. Uomoto JM, Esselman PC. Traumatic brain injury and chronic pain: differential types and rates by head injury severity. Arch Phys Med Rehabil 1993;74(1):61–4

5. Romesser J, Booth J, Benge J, et al. Mild traumatic brain injury and pain in Operation Iraqi Freedom/Operation Enduring Freedom veterans. J Rehabil Res Dev 2012;49(7):1127–36

6. Herrero Babiloni A, Bouferguene Y, Exposto FG, et al. The prevalence of persistent post-traumatic headache in adult civilian traumatic brain injury: a systematic review and meta-analysis on the past 14 years. Pain 2023;164(12):2627–2641, doi:10.1097/j.pain.0000000000002949

7. Ashina H, Iljazi A, Al-Khazali HM, et al. Persistent post-traumatic headache attributed to mild traumatic brain injury: Deep phenotyping and treatment patterns. Cephalalgia 2020;40(6):554–564, doi:10.1177/0333102420909865

8. Shahidi B, Bursch RW, Carmel JS, et al. Greater Severity and Functional Impact of Post-traumatic Headache in Veterans With Comorbid Neck Pain Following Traumatic Brain Injury. Mil Med 2021;186(11-12):1207–1214, doi:10.1093/milmed/usaa532

9. Brown AW, Watanabe TK, Hoffman JM, et al. Headache after traumatic brain injury: a national survey of clinical practices and treatment approaches. PM R 2015;7(1):3–8, doi:10.1016/j.pmrj.2014.06.016

10. Khoury S, Benavides R. Pain with traumatic brain injury and psychological disorders. Prog Neuropsychopharmacol Biol Psychiatry 2017, doi:10.1016/j.pnpbp.2017.06.007

11. Watanabe TK, Bell KR, Walker WC, et al. Systematic review of interventions for post-traumatic headache. PM R 2012;4(2):129–40, doi:10.1016/j.pmrj.2011.06.003

12. Lucas S. Posttraumatic Headache: Clinical Characterization and Management. Curr Pain Headache Rep 2015;19(10):48, doi:10.1007/s11916-015-0520-1

13. Ashina H, Porreca F, Anderson T, et al. Post-traumatic headache: epidemiology and pathophysiological insights. Nat Rev Neurol 2019;15(10):607–617, doi:10.1038/s41582-019-0243-8

14. Ruff RL, Blake K. Pathophysiological links between traumatic brain injury and post-traumatic headaches. F1000Res 2016;5(doi:10.12688/f1000research.9017.1

15. Monteith TS, Borsook D. Insights and advances in post-traumatic headache: research considerations. Curr Neurol Neurosci Rep 2014;14(2):428, doi:10.1007/s11910-013-0428-2

16. Lucas S, Hoffman JM, Bell KR, et al. A prospective study of prevalence and characterization of headache following mild traumatic brain injury. Cephalalgia 2014;34(2):93–102, doi:10.1177/0333102413499645

17. Mavroudis I, Ciobica A, Luca AC, et al. Post-Traumatic Headache: A Review of Prevalence, Clinical Features, Risk Factors, and Treatment Strategies. J Clin Med 2023;12(13), doi:10.3390/jcm12134233

18. Iyengar S, Johnson KW, Ossipov MH, et al. CGRP and the Trigeminal System in Migraine. Headache 2019;59(5):659–681, doi:10.1111/head.13529

19. Jensen R. Peripheral and central mechanisms in tension-type headache: an update. Cephalalgia 2003;23 Suppl 1(49-52, doi:10.1046/j.1468-2982.2003.00574.x

20. Leung A. Addressing chronic persistent headaches after MTBI as a neuropathic pain state. J Headache Pain 2020;21(1):77, doi:10.1186/s10194-020-01133-2

21. Ashina H, Al-Khazali HM, Iljazi A, et al. Total tenderness score and pressure pain thresholds in persistent post-traumatic headache attributed to mild traumatic brain injury. J Headache Pain 2022;23(1):96, doi:10.1186/s10194-022-01457-1

22. Defrin R, Gruener H, Schreiber S, et al. Quantitative somatosensory testing of subjects with chronic post-traumatic headache: implications on its mechanisms. Eur J Pain 2010;14(9):924–31, doi:10.1016/j.ejpain.2010.03.004

23. Defrin R, Riabinin M, Feingold Y, et al. Deficient pain modulatory systems in patients with mild traumatic brain and chronic post-traumatic headache: implications for its mechanism. J Neurotrauma 2015;32(1):28–37, doi:10.1089/neu.2014.3359

24. Levy D, Gruener H, Riabinin M, et al. Different clinical phenotypes of persistent post-traumatic headache exhibit distinct sensory profiles. Cephalalgia 2020;40(7):675–688, doi:10.1177/0333102419896368

25. Chong CD, Nikolova J, Dumkrieger GM. Migraine and Posttraumatic Headache: Similarities and Differences in Brain Network Connectivity. Semin Neurol 2022;42(4):441–448, doi:10.1055/s-0042-1757929

26. Schwedt TJ. Structural and Functional Brain Alterations in Post-traumatic Headache Attributed to Mild Traumatic Brain Injury: A Narrative Review. Front Neurol 2019;10(615, doi:10.3389/fneur.2019.00615

27. Smitha KA, Akhil Raja K, Arun KM, et al. Resting state fMRI: A review on methods in resting state connectivity analysis and resting state networks. Neuroradiol J 2017;30(4):305–317, doi:10.1177/1971400917697342

28. Bushnell MC, Ceko M, Low LA. Cognitive and emotional control of pain and its disruption in chronic pain. Nat Rev Neurosci 2013;14(7):502–11, doi:10.1038/nrn3516

29. De Ridder D, Vanneste S, Smith M, et al. Pain and the Triple Network Model. Front Neurol 2022;13(757241, doi:10.3389/fneur.2022.757241

30. Niu X, Bai L, Sun Y, et al. Disruption of periaqueductal grey-default mode network functional connectivity predicts persistent post-traumatic headache in mild traumatic brain injury. J Neurol Neurosurg Psychiatry 2019;90(3):326–332, doi:10.1136/jnnp-2018-318886

31. Bosak N, Branco P, Kuperman P, et al. Brain Connectivity Predicts Chronic Pain in Acute Mild Traumatic Brain Injury. Ann Neurol 2022;92(5):819–833, doi:10.1002/ana.26463

32. Jang SH, Park SM, Kwon HG. Relation between injury of the periaqueductal gray and central pain in patients with mild traumatic brain injury: Observational study. Medicine (Baltimore) 2016;95(26):e4017, doi:10.1097/MD.0000000000004017

33. Ofoghi Z, Dewey D, Barlow KM. A Systematic Review of Structural and Functional Imaging Correlates of Headache or Pain after Mild Traumatic Brain Injury. J Neurotrauma 2020;37(7):907–923, doi:10.1089/neu.2019.6750

34. van der Horn HJ, Liemburg EJ, Aleman A, et al. Brain Networks Subserving Emotion Regulation and Adaptation after Mild Traumatic Brain Injury. J Neurotrauma 2016;33(1):1–9, doi:10.1089/neu.2015.3905

35. Bartsch T, Goadsby PJ. The trigeminocervical complex and migraine: current concepts and synthesis. Curr Pain Headache Rep 2003;7(5):371–6

36. Bogduk N. Cervicogenic headache: anatomic basis and pathophysiologic mechanisms. Curr Pain Headache Rep 2001;5(4):382–6

37. Gironda RJ, Clark ME, Ruff RL, et al. Traumatic brain injury, polytrauma, and pain: challenges and treatment strategies for the polytrauma rehabilitation. Rehabil Psychol 2009;54(3):247–58, doi:10.1037/a0016906

38. Caberwal T, Cecchini AS, Wentz LM, et al. Prevalence of Neck Pain in Soldiers as a Result of Mild Traumatic Brain Injury-Associated Trauma. Mil Med 2024;189(1-2):e182–e187, doi:10.1093/milmed/usad228

39. Packard RC. The relationship of neck injury and post-traumatic headache. Curr Pain Headache Rep 2002;6(4):301–7, doi:10.1007/s11916-002-0051-4

40. Gil C, Decq P. How similar are whiplash and mild traumatic brain injury? A systematic review. Neurochirurgie 2021;67(3):238–243, doi:10.1016/j.neuchi.2021.01.016

41. Fakhran S, Qu C, Alhilali LM. Effect of the Suboccipital Musculature on Symptom Severity and Recovery after Mild Traumatic Brain Injury. AJNR Am J Neuroradiol 2016;37(8):1556–60, doi:10.3174/ajnr.A4730

42. Elkin BS, Elliott JM, Siegmund GP. Whiplash Injury or Concussion? A Possible Biomechanical Explanation for Concussion Symptoms in Some Individuals Following a Rear-End Collision. J Orthop Sports Phys Ther 2016;46(10):874–885, doi:10.2519/jospt.2016.7049

43. Evans RW. Persistent post-traumatic headache, postconcussion syndrome, and whiplash injuries: the evidence for a non-traumatic basis with an historical review. Headache 2010;50(4):716–24, doi:10.1111/j.1526-4610.2010.01645.x

44. Elliott JM. Are there implications for morphological changes in neck muscles after whiplash injury? Spine (Phila Pa 1976) 2011;36(25 Suppl):S205-10, doi:10.1097/BRS.0b013e3182387f57

45. Elliott JM, Pedler AR, Jull GA, et al. Differential changes in muscle composition exist in traumatic and nontraumatic neck pain. Spine (Phila Pa 1976) 2014;39(1):39-47, doi:10.1097/BRS.0000000000000033

46. Fleckenstein JL, Watumull D, Conner KE, et al. Denervated human skeletal muscle: MR imaging evaluation. Radiology 1993;187(1):213–8, doi:10.1148/radiology.187.1.8451416

47. Ashina H, Eigenbrodt AK, Seifert T, et al. Post-traumatic headache attributed to traumatic brain injury: classification, clinical characteristics, and treatment. Lancet Neurol 2021;20(6):460–469, doi:10.1016/S1474-4422(21)00094-6

48. Maleki N, Finkel A, Cai G, et al. Post-traumatic Headache and Mild Traumatic Brain Injury: Brain Networks and Connectivity. Curr Pain Headache Rep 2021;25(3):20, doi:10.1007/s11916-020-00935-y

49. Management of Concussion/m TBIWG. VA/DoD Clinical Practice Guideline for Management of Concussion/Mild Traumatic Brain Injury. J Rehabil Res Dev 2009;46(6):CP1-68

50. Headache Classification Committee of the International Headache Society (IHS) The International Classification of Headache Disorders, 3rd edition. Cephalalgia 2018;38(1):1-211, doi:10.1177/0333102417738202

51. Crofford LJ. Adverse effects of chronic opioid therapy for chronic musculoskeletal pain. Nat Rev Rheumatol 2010;6(4):191–7, doi:10.1038/nrrheum.2010.24

52. Fortier CB, Amick MM, Grande L, et al. The Boston Assessment of Traumatic Brain Injury-Lifetime (BAT-L) semistructured interview: evidence of research utility and validity. J Head Trauma Rehabil 2014;29(1):89–98, doi:10.1097/HTR.0b013e3182865859

53. Kim S, Currao A, Fonda JR, et al. Diagnostic Accuracy of the Boston Assessment of Traumatic Brain Injury-Lifetime Clinical Interview Compared to Department of Defense Medical Records. Mil Med 2023;188(11-12):3561–3569, doi:10.1093/milmed/usac162

54. Castien RF, Blankenstein AH, Windt DA, et al. Minimal clinically important change on the Headache Impact Test-6 questionnaire in patients with chronic tension-type headache. Cephalalgia 2012;32(9):710–4, doi:10.1177/0333102412449933

55. Yang M, Rendas-Baum R, Varon SF, et al. Validation of the Headache Impact Test (HIT-6) across episodic and chronic migraine. Cephalalgia 2011;31(3):357–67, doi:10.1177/0333102410379890

56. Moses MJ, Tishelman JC, Stekas N, et al. Comparison of Patient Reported Outcome Measurement Information System With Neck Disability Index and Visual Analog Scale in Patients With Neck Pain. Spine (Phila Pa 1976) 2019;44(3):E162-E167, doi:10.1097/BRS.0000000000002796

57. Pilkonis PA, Choi SW, Reise SP, et al. Item banks for measuring emotional distress from the Patient-Reported Outcomes Measurement Information System (PROMIS(R)): depression, anxiety, and anger. Assessment 2011;18(3):263–83, doi:10.1177/1073191111411667

58. Keane TM, Fairbank JA, Caddell JM, et al. Clinical evaluation of a measure to assess combat exposure. Psychol Assess 1989;1(1):53–55

59. Bovin MJ, Marx BP, Weathers FW, et al. Psychometric properties of the PTSD Checklist for Diagnostic and Statistical Manual of Mental Disorders-Fifth Edition (PCL-5) in veterans. Psychol Assess 2016;28(11):1379–1391, doi:10.1037/pas0000254

60. McCracken LM, Dhingra L. A short version of the Pain Anxiety Symptoms Scale (PASS-20): preliminary development and validity. Pain Res Manag 2002;7(1):45–50, doi:10.1155/2002/517163

61. Rogers AH, Gallagher MW, Garey L, et al. Pain Anxiety Symptoms Scale-20: An empirical evaluation of measurement invariance across race/ethnicity, sex, and pain. Psychol Assess 2020;32(9):818–828, doi:10.1037/pas0000884

62. Walton DM, Wideman TH, Sullivan MJ. A Rasch analysis of the pain catastrophizing scale supports its use as an interval-level measure. Clin J Pain 2013;29(6):499–506, doi:10.1097/AJP.0b013e318269569c

63. Nicholas MK. The pain self-efficacy questionnaire: Taking pain into account. Eur J Pain 2007;11(2):153–63, doi:10.1016/j.ejpain.2005.12.008

64. Mainero C, Boshyan J, Hadjikhani N. Altered functional magnetic resonance imaging resting-state connectivity in periaqueductal gray networks in migraine. Ann Neurol 2011;70(5):838–45, doi:10.1002/ana.22537

65. Coulombe MA, Erpelding N, Kucyi A, et al. Intrinsic functional connectivity of periaqueductal gray subregions in humans. Hum Brain Mapp 2016;37(4):1514–30, doi:10.1002/hbm.23117

66. Amin FM, Hougaard A, Magon S, et al. Altered thalamic connectivity during spontaneous attacks of migraine without aura: A resting-state fMRI study. Cephalalgia 2018;38(7):1237–1244, doi:10.1177/0333102417729113

67. Hemington KS, Wu Q, Kucyi A, et al. Abnormal cross-network functional connectivity in chronic pain and its association with clinical symptoms. Brain Struct Funct 2016;221(8):4203–4219, doi:10.1007/s00429-015-1161-1

68. Vassal M, Charroud C, Deverdun J, et al. Recovery of functional connectivity of the sensorimotor network after surgery for diffuse low-grade gliomas involving the supplementary motor area. J Neurosurg 2017;126(4):1181–1190, doi:10.3171/2016.4.JNS152484

69. Reeder SB, Pineda AR, Wen Z, et al. Iterative decomposition of water and fat with echo asymmetry and least-squares estimation (IDEAL): application with fast spin-echo imaging. Magn Reson Med 2005;54(3):636–44, doi:10.1002/mrm.20624

70. Shahidi B, Parra CL, Berry DB, et al. Contribution of Lumbar Spine Pathology and age to Paraspinal Muscle Size and fatty Infiltration. Spine (Phila Pa 1976) 2016, doi:10.1097/BRS.0000000000001848

71. Fortin M, Battié MC. Quantitative paraspinal muscle measurements: inter-software reliability and agreement using OsiriX and ImageJ. Phys Ther 2012;92(6):853–64, doi:10.2522/ptj.20110380

72. Rosset A, Spadola L, Ratib O. OsiriX: an open-source software for navigating in multidimensional DICOM images. J Digit Imaging 2004;17(3):205–16, doi:10.1007/s10278-004-1014-6

73. Berry DB, Shahidi B, Rodriguez-Soto A, et al. Lumbar muscle structure predicts operational postures in active-duty Marines. JOSPT 2018 (In Press);

74. Carey C, Saxe J, White FA, et al. An Exploratory Study of Endogenous Pain Modulatory Function in Patients Following Mild Traumatic Brain Injury. Pain Med 2019;20(11):2198–2207, doi:10.1093/pm/pnz058

75. Young IA, Dunning J, Butts R, et al. Psychometric properties of the Numeric Pain Rating Scale and Neck Disability Index in patients with cervicogenic headache. Cephalalgia 2019;39(1):44–51, doi:10.1177/0333102418772584

76. Young Ia Pt D, Dunning J Pt DPT, Butts R Pt P, et al. Reliability, construct validity, and responsiveness of the neck disability index and numeric pain rating scale in patients with mechanical neck pain without upper extremity symptoms. Physiother Theory Pract 2019;35(12):1328-1335, doi:10.1080/09593985.2018.1471763

77. Arendt-Nielsen L, Petersen-Felix S. Wind-up and neuroplasticity: is there a correlation to clinical pain? Eur J Anaesthesiol Suppl 1995;10(1-7

78. Cathcart S, Winefield AH, Rolan P, et al. Reliability of temporal summation and diffuse noxious inhibitory control. Pain Res Manag 2009;14(6):433–8, doi:10.1155/2009/523098

79. Youssef AM, Macefield VG, Henderson LA. Pain inhibits pain; human brainstem mechanisms. Neuroimage 2016;124(Pt A):54–62, doi:10.1016/j.neuroimage.2015.08.060

80. Yarnitsky D, Bouhassira D, Drewes AM, et al. Recommendations on practice of conditioned pain modulation (CPM) testing. Eur J Pain 2015;19(6):805–6, doi:10.1002/ejp.605

81. Costa YM, Morita-Neto O, de Araujo-Junior EN, et al. Test-retest reliability of quantitative sensory testing for mechanical somatosensory and pain modulation assessment of masticatory structures. J Oral Rehabil 2017;44(3):197–204, doi:10.1111/joor.12477

82. Coulombe-Leveque A, Tousignant-Laflamme Y, Leonard G, et al. The effect of conditioning stimulus intensity on conditioned pain modulation (CPM) hypoalgesia. Can J Pain 2021;5(1):22–29, doi:10.1080/24740527.2020.1855972

83. Lewis GN, Heales L, Rice DA, et al. Reliability of the conditioned pain modulation paradigm to assess endogenous inhibitory pain pathways. Pain Res Manag 2012;17(2):98–102

84. Mitrovic K, Petrusic I, Radojicic A, et al. Migraine with aura detection and subtype classification using machine learning algorithms and morphometric magnetic resonance imaging data. Front Neurol 2023;14(1106612, doi:10.3389/fneur.2023.1106612

85. Breiman L. Random Forests. Machine Learning 2001;45(1):5-32, doi:10.1023/A:1010933404324

86. Rousseeuw PJ. Silhouettes: a graphical aid to the interpretation and validation of cluster analysis. J Comput Appl Math 1987;20(53-65

87. Team RC. R: A language and environment for statistical computing. 2021. Available from: https://www.R-project.org/.

88. Linnman C, Moulton EA, Barmettler G, et al. Neuroimaging of the periaqueductal gray: state of the field. Neuroimage 2012;60(1):505–22, doi:10.1016/j.neuroimage.2011.11.095

89. Silvestro M, Tessitore A, Caiazzo G, et al. Disconnectome of the migraine brain: a "connectopathy" model. J Headache Pain 2021;22(1):102, doi:10.1186/s10194-021-01315-6

90. Vogt BA. Midcingulate cortex: Structure, connections, homologies, functions and diseases. J Chem Neuroanat 2016;74(28-46, doi:10.1016/j.jchemneu.2016.01.010

91. Vogt BA. Cingulate neurobiology and disease. Oxford University Press: New York, NY; 2009.

92. Vogt BA, Berger GR, Derbyshire SW. Structural and functional dichotomy of human midcingulate cortex. Eur J Neurosci 2003;18(11):3134–44, doi:10.1111/j.1460-9568.2003.03034.x

93. Kucyi A, Hodaie M, Davis KD. Lateralization in intrinsic functional connectivity of the temporoparietal junction with salience- and attention-related brain networks. J Neurophysiol 2012;108(12):3382–92, doi:10.1152/jn.00674.2012

94. Shpaner M, Kelly C, Lieberman G, et al. Unlearning chronic pain: A randomized controlled trial to investigate changes in intrinsic brain connectivity following Cognitive Behavioral Therapy. Neuroimage Clin 2014;5(365-76, doi:10.1016/j.nicl.2014.07.008

95. Lim M, Kim DJ, Nascimento TD, et al. High-definition tDCS over primary motor cortex modulates brain signal variability and functional connectivity in episodic migraine. Clin Neurophysiol 2024;161(101-111, doi:10.1016/j.clinph.2024.02.012

96. Kong J, Tu PC, Zyloney C, et al. Intrinsic functional connectivity of the periaqueductal gray, a resting fMRI study. Behav Brain Res 2010;211(2):215–9, doi:10.1016/j.bbr.2010.03.042

97. Murphy K, Birn RM, Handwerker DA, et al. The impact of global signal regression on resting state correlations: are anti-correlated networks introduced? Neuroimage 2009;44(3):893–905, doi:10.1016/j.neuroimage.2008.09.036

98. Carbonell F, Bellec P, Shmuel A. Quantification of the impact of a confounding variable on functional connectivity confirms anti-correlated networks in the resting-state. Neuroimage 2014;86(343-53, doi:10.1016/j.neuroimage.2013.10.013

99. Liu Y, Huang L, Li M, et al. Anticorrelated networks in resting-state fMRI-BOLD data. Biomed Mater Eng 2015;26 Suppl 1(S1201-11, doi:10.3233/BME-151417

100. Andersen S, Petersen MW, Svendsen AS, et al. Pressure pain thresholds assessed over temporalis, masseter, and frontalis muscles in healthy individuals, patients with tension-type headache, and those with migraine--a systematic review. Pain 2015;156(8):1409–1423, doi:10.1097/j.pain.0000000000000219

101. Castien RF, van der Wouden JC, De Hertogh W. Pressure pain thresholds over the cranio-cervical region in headache: a systematic review and meta-analysis. J Headache Pain 2018;19(1):9, doi:10.1186/s10194-018-0833-7

102. Nahman-Averbuch H, Callahan D, Darken R, et al. Harnessing the conditioned pain modulation response in migraine diagnosis, outcome prediction, and treatment-A narrative review. Headache 2023;63(8):1167–1177, doi:10.1111/head.14601

103. Nahman-Averbuch H, Granovsky Y, Coghill RC, et al. Waning of "conditioned pain modulation": a novel expression of subtle pronociception in migraine. Headache 2013;53(7):1104–15, doi:10.1111/head.12117

104. Sterling M, Elliott JM, Cabot PJ. The course of serum inflammatory biomarkers following whiplash injury and their relationship to sensory and muscle measures: a longitudinal cohort study. PLoS One 2013;8(10):e77903, doi:10.1371/journal.pone.0077903

105. 105. Flaores AG, Ferisgan M, Onita D, et al. The smallest sample size for the desired diagnosis accuracy. 2017. Available from: https://www.researchgate.net/publication/319422745 [Last Accessed; July 12].

106. Peterlin BL, Nijjar SS, Tietjen GE. Post-traumatic stress disorder and migraine: epidemiology, sex differences, and potential mechanisms. Headache 2011;51(6):860–8, doi:10.1111/j.1526-4610.2011.01907.x

107. Knackstedt H, Krakenes J, Bansevicius D, et al. Magnetic resonance imaging of craniovertebral structures: clinical significance in cervicogenic headaches. J Headache Pain 2012;13(1):39–44, doi:10.1007/s10194-011-0387-4

108. Li Q, Shen H, Li M. Magnetic resonance imaging signal changes of alar and transverse ligaments not correlated with whiplash-associated disorders: a meta-analysis of case-control studies. Eur Spine J 2013;22(1):14–20, doi:10.1007/s00586-012-2490-x

109. Dumkrieger G, Chong CD, Ross K, et al. Static and dynamic functional connectivity differences between migraine and persistent post-traumatic headache: A resting-state magnetic resonance imaging study. Cephalalgia 2019;39(11):1366–1381, doi:10.1177/0333102419847728

