## Appendix A for "Biological markers of brain network connectivity and pain sensitivity distinguish low coping from high coping Veterans with persistent post-traumatic headache"

**Appendix A. Data processing methods for resting state fMRI**

The first two TRs were removed from all resting state runs using `3dTcat`. Outliers and potential censoring were identified for each run and a base registration volume was identified across runs that met the minimum outlier volume for each subject. We applied `3dDespike` to each run and aligned slice timing with `3dTshift`. Anatomical alignment to the EPI registration base was computed with `align_epi_anat.py`, and each volume in the timeseries was registered to the base image using `3dvolreg`. A mask was created to define the extent of the warp, and transformations were applied with `3dNwarpApply`. The mean and intersection masks were computed and applied to the EPI data. EPI was aligned with the anatomical dataset with `3dAllineate`.

Next, anatomical datasets were warped non-linearly, and each volume was blurred with `3dmerge`. A union mask of EPI datasets was created, and the anatomical dataset was resampled. Masks were generated for CSF, GM, and WM using `3dmask_tool`, and ROI masks for the ventricles were computed for each run. The top 3 signal components were created to account for signal noise for each ventricle mask. Outlier and motion censor files were combined, and a bandpass filter was applied (0.01-0.1 Hz). The primary region of interest (ROI) for the PAG was defined by a 3-mm radius sphere around MNI coordinates x,y,z (2,-30,-6).

Deconvolution matrices were created in `3dDeconvolve` for each seed in addition to regressors of non-interest for each run: three components from ventricle masks, linear motion (x,y,z), derivative of motion, baseline trend, linear trend, and quadratic trend. The `3dTproject` tool was used to project out the regression matrix, and the X-matrix was checked for large pairwise correlations and degrees of freedom. All runs were concatenated, and the ANATICOR result was generated by projecting out voxel-wise regressors. Temporal signal-to-noise ratio datasets were computed, and global correlation average (GCOR) was calculated but global signal was not removed from the model. Pearson correlations between the PAG and each network seed four-dimensional residuals and the average seed time-series were computed. Correlation coefficients were converted to Z scores with Fisher’s r-to-z transform. Extraction ROIs were selected within the *default mode network* (DMN: medial prefrontal cortex (mPFC ±2, 56, 16), posterior cingulate cortex/precuneus (PCC 8,-50,28), subgenual anterior cingulate cortex (sgACC ±5,34,-4), lateral temporal cortex (right LTC 56,-14,-20, left LTC -56,-20,-16), inferior parietal cortex (right IPC 50,-64,30, left IPC -44,-68, 28)), *salience network* (SN: temporoparietal junction (right TPJ 62,-36,28, left TPJ -60,-38,26), anterior insula (right aINS 34,18,4 left aINS -34,8,6), dorsolateral prefrontal cortex (right DLPFC 34,46,22, left DLPFC -38,40,28), middle cingulate cortex (MCC 2,12,34), and *sensorimotor network* (SMN: primary motor cortex (right M1 40,-20,54, left M1 -43,-18,52), primary sensory cortex (right S1 48,-16,38, left S1 -49,-19,36), supplementary motor area (SMA ±2,3,53), thalamus (right 22,-24,0, left -22,-28,6), posterior insula (right pINS 39,-4,10, left pINS -39,2,9), cerebellum (±46,-58,-30)).

Finally, blur was estimated using `3dFWHMx` for each run and blur across runs was estimated, with results recorded in the `blur_est.$subj.1D` file. All steps were performed within the context of AFNI processing tools, ensuring that the data were prepared and cleaned for further analysis.
